# A Novel Evidence-Based Predictor Tool for Hospitalization and Length of Stay: Insights from COVID19 Patients in New York City

**DOI:** 10.1101/2021.04.24.21256042

**Authors:** Maan El Halabi, James Feghali, Paulino Tallón de Lara, Bharat Narasimhan, Kam Ho, Joseph Saabiye, Judy Huang, Georgina Osorio, Joseph Mathew, Juan Wisnivesky, David Steiger

**Author notes:** These authors contributed equally to this manuscript (Maan El Halabi, James Feghali). **Address for correspondence:** David J Steiger, MD, Division of Pulmonary and Critical Care Medicine Icahn School of Medicine at Mount Sinai New York, NY 10019, USA; **E-mail:**.

## Abstract

**Background:** Coronavirus disease 2019 (COVID-19) has evolved into a true global pandemic infecting more than 30 million people worldwide. Predictive models for key outcomes have the potential to optimize resource utilization and patient outcome as outbreaks continue to occur worldwide. We aimed to design and internally validate a web-based calculator predictive of hospitalization and length of stay (LOS) in a large cohort of COVID-19 positive patients presenting to the Emergency Department (ED) in a New York City health system.

**Methods:** The study cohort consisted of consecutive adult (>18 years) patients presenting to the ED of one of the Mount Sinai Health System hospitals between March, 2020 and April, 2020 who were diagnosed with COVID-19. Logistic regression was utilized to construct predictive models for hospitalization and prolonged (>3 days) LOS. Discrimination was evaluated using area under the receiver operating curve (AUC). Internal validation with bootstrapping was performed, and a web-based calculator was implemented.

**Results:** The cohort consisted of 5859 patients with a hospitalization rate of 65% and a prolonged LOS rate of 75% among hospitalized patients. Independent predictors of hospitalization included older age (OR=6.29; 95% CI [1.83-2.63], >65 vs. 18-44), male sex (OR=1.35 [1.17-1.55]), chronic obstructive pulmonary disease (OR=1.74; [1.00-3.03]), hypertension (OR=1.39; [1.13-1.70]), diabetes (OR=1.45; [1.16-1.81]), chronic kidney disease (OR=1.69; [1.23-2.32]), elevated maximum temperature (OR=4.98; [4.28-5.79]), and low minimum oxygen saturation (OR=13.40; [10.59-16.96]). Predictors of extended LOS included older age (OR=1.03 [1.02-1.04], per year), chronic kidney disease (OR=1.91 [1.35-2.71]), elevated maximum temperature (OR=2.91 [2.40- 3.53]), and low minimum percent oxygen saturation (OR=3.89 [3.16-4.79]). AUCs of 0.881 and 0.770 were achieved for hospitalization and LOS, respectively. A calculator was made available under the following URL: https://covid19-outcome-prediction.shinyapps.io/COVID19_Hospitalization_Calculator/

**Conclusion:** The prediction tool derived from this study can be used to optimize resource allocation, guide quality of care, and assist in designing future studies on the triage and management of patients with COVID-19.

## Introduction

Ten months after the initial outbreak of the 2019 novel Coronavirus (SARS-Cov-2) in Wuhan, China, the disease evolved into a global pandemic with more than thirty-seven million people infected and one million deaths worldwide (Date of this data?). By October 2020, the United States had more than seven million cases with New York becoming the epicenter of the pandemic, accounting for a large proportion of infections in the United States.^1^

The scale of infection imposed a major strain on medical infrastructure and resources leading to substantial shortages in the early stages of the pandemic. Advanced age, hypoxia upon presentation, abnormal chest imaging, and elevated inflammatory markers appear to be strong predictors of worse outcomes.^2–6^ Identifying predictors of hospitalization and length of stay (LOS), which are both highly relevant outcomes in patients with COVID-19, are being described. Increasing age and multimorbidity were associated with hospitalization in two recent studies of patients who tested positive for SARS-CoV-2.^4,7^ However, additional studies are needed to validate these findings. Understanding the determinants of the need for hospitalization and the projected length of hospital stay can optimize the utilization of hospital resources including healthcare personnel, intensive care unit (ICU) beds, ventilators, and personal protective equipment (PPE). It would also provide opportunities for timely health care assessment for patients, aid in medical triaging, and improve shared decision-making.^5^ The aim of this study is to describe the demographics, clinical characteristics, and outcomes in a large COVID-19 cohort and to derive predictive models for hospitalization and prolonged LOS.

## Methods

### Patients

Data for the study was obtained from the Mount Sinai Data Warehouse, a registry of de-identified patient data extracted from the electronic medical record system (EPIC) across the Mount Sinai network. The database consisted of consecutive adult (>18 years) patients presenting to an emergency department (ED) of one of the Mount Sinai Health System hospitals: Mount Sinai Hospital, Mount Sinai Morningside, Mount Sinai West, Mount Sinai Brooklyn and Mount Sinai Queens, all located in New York City between March 20 and April 23 2020 and who were diagnosed with SARS-COV2. Diagnosis was confirmed by reverse transcriptase-polymerase-chain-reaction (RT-PCR) of nasopharyngeal or oropharyngeal specimens. This data was collected starting from the initial phases of the COVID-19 pandemic in New York City, and all tested patients presented to the ED with symptoms consistent with COVID-19 including fever, cough, diarrhea, or shortness of breath. This study was approved by the Mount Sinai Institutional Review Board (IRB). Since no direct patient contact or intervention from the study group was needed, no patient consent was required.

### Variables

Patient demographics, diagnosis codes (International Classification of Diseases-9/10- Clinical Modification (ICD-9/10-CM) code), and clinical data including symptoms, vital signs as well as laboratory data were collected on presentation. We defined a pre-existing condition as the presence of diagnosis codes (ICD 9/10) associated with specific diseases (Supplemental Table 1).The earliest available laboratory results during the first 24 hours were used in the prediction analysis.

**Table 1.** Patient demographics, comorbidities, and vital signs on presentation.

| Characteristic* | Value |
| --- | --- |
| *Demographics* |  |
| Age, yrs, mean (SD) | 60.5 ± 17.5 |
| Race (n=5669) |  |
| White | 1373 (24) |
| African American | 1576 (28) |
| Asian | 229 (4) |
| Native Hawaiian or Other Pacific Islander | 140 (2) |
| Other | 2351 (41) |
| Sex, female | 2606 (44) |
| Body mass index, kg/m<sup>2</sup> (n=4201) | 29.19 ± 8.39 |
| Ever smoked (n=4424) | 1282 (29) |
| *Comorbidities* |  |
| Asthma | 257 (4) |
| COPD | 176 (3) |
| Hypertension | 1627 (28) |
| Diabetes | 1157 (20) |
| Chronic kidney disease | 524 (9) |
| HIV | 90 (2) |
| Cancer | 322 (5) |
| *Vital Signs* |  |
| Temperature max, degrees Celsius (n=5848) | 38 ± 1.2 |
| Heart rate, beats per minute (n=5850) | 95.7 ± 19 |
| Respiratory rate, breaths per minute (n=5844) | 20.5 ± 5.3 |
| Systolic blood pressure, mmHg (n=5841) | 131.6 ± 23.1 |
| Diastolic blood pressure, mmHg (n=5841) | 75.5 ± 13.8 |
| %O<sub>2</sub> saturation minimum (n=5848) | 87.5 ± 15.6 |

HIV: Human immunodeficiency virus; COPD: Chronic obstructive pulmonary disorder
\* Numbers in parentheses represent number of patients without missing data

The primary outcome was the need for hospital admission after presentation. A secondary outcome included an extended LOS defined as hospitalization lasting more than 3 days among patients that completed follow-up (i.e., we excluded patients that were still hospitalized at the time of data analysis). As recommended by Hintz et al.,^8^ we also excluded patients who died within 3 days to avoid a potentially misleading LOS value denoting good outcome. The LOS cut-off of 3 days was selected for several reasons. Public health authorities including the Centers for Disease Control and Prevention, European Centre for Disease Prevention and Control, and the National Centre for Infectious Disease state that three days of symptom resolution, namely fever and respiratory symptoms, is the cut-off for safe discharge.^1,9^ Moreover, median time to readmission,^10^ median time to radiographic progression,^11^ as well as median time to clinical deterioration following admission^12^ were all 3 days. Patients were also tracked for mortality, need for intensive care, and intubation.

### Statistical Analysis

Analyses were performed using R (R Foundation for Statistical Computing, Vienna, Austria) with statistical significance set at *P*≤0.05. Descriptive statistics (n, % for categorical variables and mean ± SD for continuous variables) were used to summarize the baseline demographic, clinical, and laboratory variables of the study population. Continuous variables were subsequently categorized using clinically relevant cut-offs. For the first outcome of interest, a univariable analysis of factors associated with the need for hospitalization consisting of a Chi-square or Fischer exact test for categorical variables and the student’s T-test for continuous variables was conducted. Characteristics with a p-value <0.1 were subsequently entered a stepwise logistic regression model. Given the amount of missing data for laboratory values, these variables were left out of this step-wise process. Collinearity between variable pairs was evaluated using a strict variance inflation factor (VIF) cut-off of 3. Model discrimination was assessed using a receiver operating characteristic (ROC) curve analysis to obtain an area under the curve (AUC). Bootstrapping with 1000 samples with replacement was utilized to calculate an optimism-corrected AUC to check for possible overfitting.^13,14^ Calibration was evaluated by using the Hosmer-Lemeshow test with an adjusted number of subgroups to account for a large sample size in our study.^15^

In hospitalized patients that completed follow-up and survived, a similar process consisting of univariable analysis, stepwise multivariable logistic regression for factors independently associated with an extended LOS.

Finally, we developed a web-based calculator that uses the models to predict the probability of a patient requiring hospitalization and extended LOS using readily available components of the history and vital signs on first patient encounter using the Shiny package from R.

## Results

### Patients

The cohort consisted of 5859 patients with a mean age of 60.5 years (SD=17.5 years) with 3253 males (56%). Racial, ethnic groups included 1373 (24%) white, 1576 (28%) black, 229 (4%) Asian, 140 (3%) Native Hawaiian/Pacific Islander, and 2351 (41%) with other ethnicities. The baseline demographic and clinical characteristics are summarized in Table 1. Laboratory values obtained during the first 24 hours are summarized in Table 2.

**Table 2.** Lab values within first 24 hours.

| Lab* | Mean ± SD |
| --- | --- |
| ALT, U/L (n=4123) | 49.7 ± 104.6 |
| AST, U/L (n=4001) | 69.1 ± 180.7 |
| Creatinine, mg/dL (n=4575) | 1.8 ± 2.4 |
| CRP, mg/L (n=1430) | 139.8 ± 96.3 |
| D-dimer, mcg/ml fibrinogen equivalent units (n=2980) | 2.6 ± 3.2 |
| Ferritin, ng/mL (n=3250) | 1519.3 ± 2592 |
| Hemoglobin, g/dL (n=2090) | 13.0 ± 3.2 |
| LDH, U/L (n=2995) | 511.6 ± 400.6 |
| Procalcitonin, ng/mL (n=3167) | 2.2 ± 16.6 |
| Troponin, ng/mL (n=2911) | 0.3 ± 2.7 |
| WBC count, x1000 cells/mm<sup>3</sup> (n=5276) | 8.4 ± 5.4 |

ALT: Alanine aminotransferase; AST: Aspartate aminotransferase; CRP: C-reactive protein; LDH: Lactate dehydrogenase; WBC: White blood cell
\* Numbers in parentheses represent number of patients without missing data

### Univariable analysis, Hospitalization

Out of 5859 patients, 3794 (65%) were hospitalized. Demographic factors and comorbidities that were significantly associated with hospitalization on univariable analysis included older age (p<0.001), race (p<0.001), male sex (p<0.001), ever smoking (p<0.001), history of chronic obstructive pulmonary disease (p<0.001), hypertension (p<0.001), obesity (p<0.001), diabetes (p<0.001), chronic kidney disease (p<0.001), and cancer (p<0.001). With regards to vital signs, a maximum temperature of 38 degrees Celsius or more (p<0.001), systolic blood pressure <90 mmHg (p<0.001), minimum percent oxygen saturation <90% (p<0.001), and several laboratory values, including elevated C-reactive protein and ferritin, were significantly associated with hospitalization. The univariable analysis is summarized in Table 3.

**Table 3.** Univariable Predictors of need for hospitalization.

| Characteristic | No admission<br>(n=2065) | Admission<br>(n=3794) | P-value |
| --- | --- | --- | --- |
| *Demographics* |  |  |  |
| Age |  |  | <0.001† |
| 19-44 yrs | 808 (39) | 397 (10) |  |
| 45-65 yrs | 854 (41) | 1350 (36) |  |
| >65 yrs | 403 (20) | 2047 (54) |  |
| Race |  |  | 0.001† |
| White | 471 (24) | 902 (25) |  |
| Black | 608 (30) | 968 (26) |  |
| Asian | 87 (4) | 142 (4) |  |
| Native Hawaiian or Other Pacific Islander | 62 (3) | 78 (2) |  |
| Other | 775 (39) | 1576 (43) |  |
| Sex, female | 1038 (50) | 1568 (41) | <0.001† |
| *Comorbidities* |  |  |  |
| Ever smoked | 331 (22) | 951 (33) | <0.001† |
| Asthma | 77 (4) | 180 (5) | 0.070 |
| COPD | 19 (1) | 157 (4) | <0.001† |
| Hypertension | 303 (15) | 1324 (35) | <0.001† |
| Obesity (BMI≥30 kg/m<sup>2</sup>) | 286 (35) | 1233 (37) | 0.306 |
| Diabetes | 210 (10) | 947 (25) | <0.001† |
| Chronic kidney disease | 72 (3) | 452 (12) | <0.001† |
| HIV | 25 (1) | 65 (2) | 0.135 |
| Cancer | 71 (3) | 251 (7) | <0.001† |
| *Vital Signs* |  |  |  |
| Temperature maximum |  |  | <0.001† |
| <38 degrees Celsius | 1638 (80) | 1447 (38) |  |
| ≥38 degrees Celsius | 416 (20) | 2347 (62) |  |
| Systolic blood pressure |  |  | <0.001† |
| <90 mmHg | 23 (1) | 99 (3) |  |
| ≥90 mmHg | 2030 (99) | 3687 (97) |  |
| %O<sub>2</sub> saturation minimum |  |  | <0.001† |
| <90 | 85 (4) | 1980 (53) |  |
| ≥90 | 1956 (96) | 1768 (47) |  |
| *Lab Values* |  |  |  |
| ALT (U/L) |  |  | 0.291 |
| 1-45* | 406 (73) | 2493 (70) |  |
| 46-135 | 128 (23) | 886 (25) |  |
| >135 | 23 (4) | 186 (5) |  |
| AST (U/L) |  |  | <0.001† |
| 1-35* | 232 (45) | 1248 (36) |  |
| 36-105 | 247 (48) | 1808 (52) |  |
| >105 | 40 (8) | 426 (12) |  |
| Creatinine (mg/dL) |  |  | <0.001† |
| 0-1* | 457 (57) | 1750 (46) |  |
| 1-2 | 273 (34) | 1258 (33) |  |
| >2 | 67 (8) | 770 (20) |  |
| CRP (mg/L) |  |  | <0.001† |
| 0.0-5.0* | 6 (5) | 23 (2) |  |
| 5.1-100 | 64 (52) | 500 (38) |  |
| >100 | 52 (43) | 784 (60) |  |
| D-dimer (mcg/ml) |  |  | 0.382 |
| 0.0-0.5* | 94 (9) | 190 (10) |  |
| 0.6-1.5 | 447 (42) | 772 (40) |  |
| >1.5 | 511 (49) | 966 (50) |  |
| Ferritin (ng/mL) |  |  | 0.005† |
| 30-400* | 370 (33) | 566 (27) |  |
| 401-500 | 81 (7) | 145 (7) |  |
| >500 | 678 (60) | 1356 (66) |  |
| Anemic hemoglobin‡ | 268 (37) | 537 (39) | 0.378 |
| LDH (U/L) |  |  | 0.243 |
| 100-220* | 65 (6) | 96 (5) |  |
| 221-250 | 50 (5) | 79 (4) |  |
| >250 | 940 (89) | 1764 (91) |  |
| Procalcitonin>0.5 ng/mL | 328 (29) | 595 (29) | 0.951 |
| Troponin (ng/mL) |  |  | 0.747 |
| 0.0-0.03* | 531 (52) | 952 (51) |  |
| 0.04-0.1 | 250 (24) | 448 (24) |  |
| >0.1 | 250 (24) | 480 (25) |  |
| WBC count (cells/mm<sup>3</sup>) |  |  | 0.904 |
| <4,000 | 156 (9) | 220 (9) |  |
| 4000-10,999 | 1158 (70) | 2534 (70) |  |
| ≥11,000 | 340 (21) | 758 (21) |  |

ALT: Alanine aminotransferase; AST: Aspartate aminotransferase; COPD: Chronic obstructive pulmonary disorder; CRP: C-reactive protein; FEU: fibrinogen equivalent units; HIV: Human immunodeficiency virus; LDH: Lactate dehydrogenase; WBC: White blood cell
\*Numbers in parentheses represent number of patients without missing data
†Statistically significant ( $P \leq 0.05$ )
‡Defined as below 13 g/dL in males and below 12 g/dL in females
\*Reference value

### Adjusted analysis, Hospitalization

Independent predictors of hospitalization included older age, male sex, chronic obstructive pulmonary disease, hypertension, diabetes, chronic kidney disease, elevated maximum temperature, and low minimum percent oxygen saturation. The optimal multivariable model resulting from stepwise logistic regression is summarized in Table 4, and consisted of age (OR=6.29; 95% CI [1.83-2.63] for older adults(>65 yrs) compared to younger adults (18-44 yrs)), male sex (OR=1.35 [1.17-1.55]), COPD (OR=1.74 [1.00-3.03]), hypertension (OR=1.39 [1.13-1.70]), diabetes (OR=1.45 [1.16-1.81]), chronic kidney disease (OR=1.69 [1.23- 2.32]), elevated maximum temperature (OR=4.98 [4.28-5.79]), and low minimum oxygen saturation (OR=13.40 [10.59-16.96]). The AUC for the model was 0.881 (95% CI: 0.872-0.890) (Figure 1). Bootstrap validation yielded negligible optimism of 0.0013 which translates into an optimism-corrected AUC of 0.880, indicating absence of significant overfitting. The Hosmer-Lemeshow test yielded a non-significant p-value, indicating appropriate calibration.

**Table 4.** Optimal multivariable logistic regression model predictive of need for hospitalization (n=5787).

| Variable | Odds ratio [95% CI] |
| --- | --- |
| Age, 18-44 years as reference |  |
| 45-65 | 2.19 [1.83-2.63] |
| >65 | 6.29 [5.15-7.69] |
| Sex, male vs. female | 1.35 [1.17-1.55] |
| COPD | 1.74 [1.00-3.03] |
| Hypertension | 1.39 [1.13-1.70] |
| Diabetes | 1.45 [1.16-1.81] |
| Chronic kidney disease | 1.69 [1.23-2.32] |
| Temperature max ≥ 38 degrees Celsius | 4.98 [4.28-5.79] |
| %O<sub>2</sub> saturation minimum < 90 | 13.40 [10.59-16.96] |

COPD: Chronic obstructive pulmonary disorder
\*Statistically significant ( $P \leq 0.05$ )
1.2% of the data were missing at least one of the above variables

**Figure 1.**
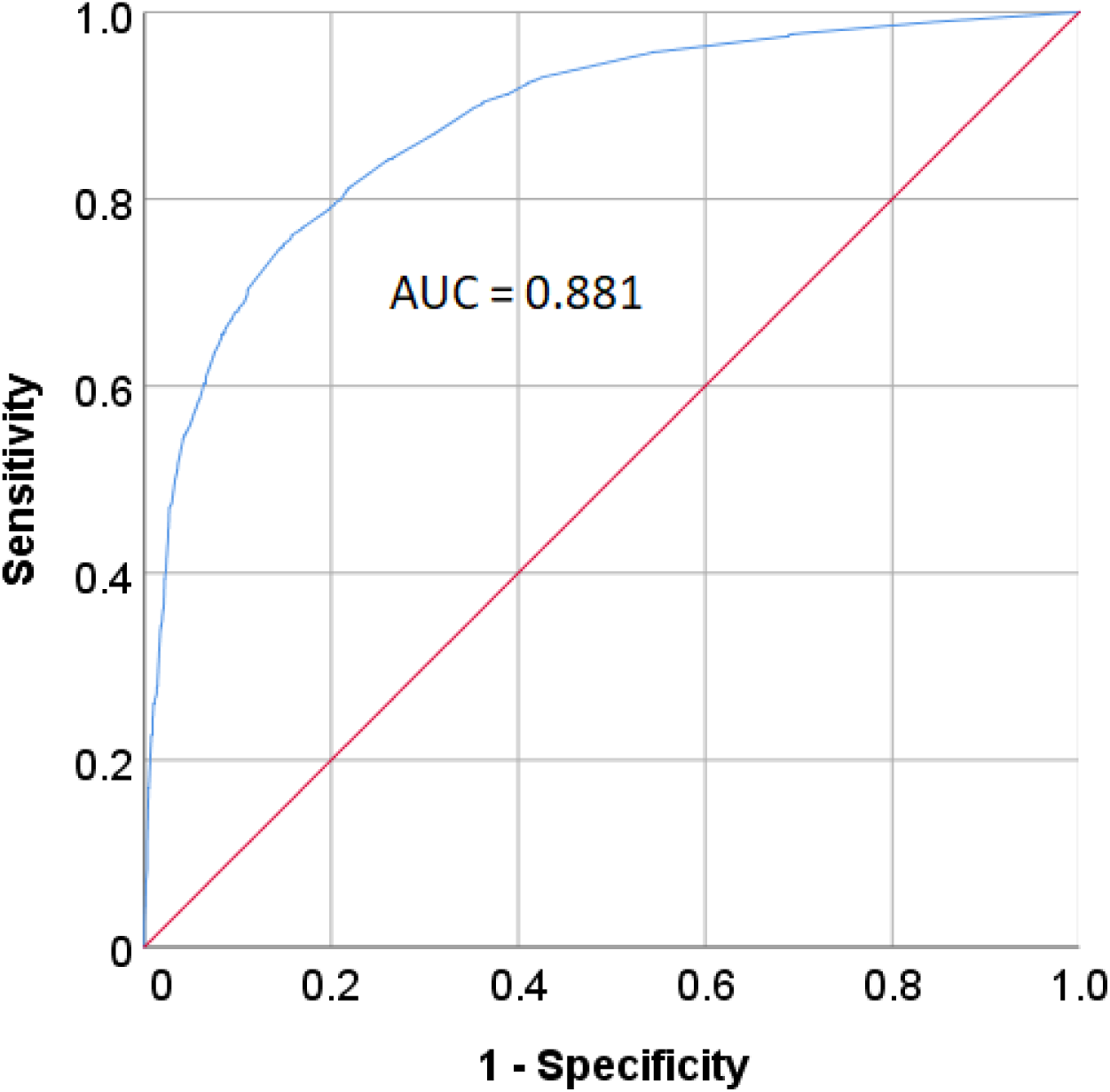
Receiver operating curve of optimal multivariable model predicting need for hospitalization with area under the curve (AUC).

### Length of stay

A flow diagram describing the course of the study population can be found in Figure 2. Out of 3794 patients requiring hospitalization, 631 (17%) were still hospitalized at the time of analysis and are still being actively followed up. These patients were excluded from subsequent analyses, leaving 3163 patients. There was a mortality rate of 28% (897/3163) among hospitalized patients and 17% (897/5228) among all patients who completed follow-up (i.e., discharged from ED or inpatient care). In addition, 16% (492/3163) of hospitalized patients required admission into an intensive care unit, and 13% (401/3163) required intubation. Among the 897 patients who died, 386 (43%) required admission into an ICU, and 315 (35%) were intubated.

**Figure 2.**
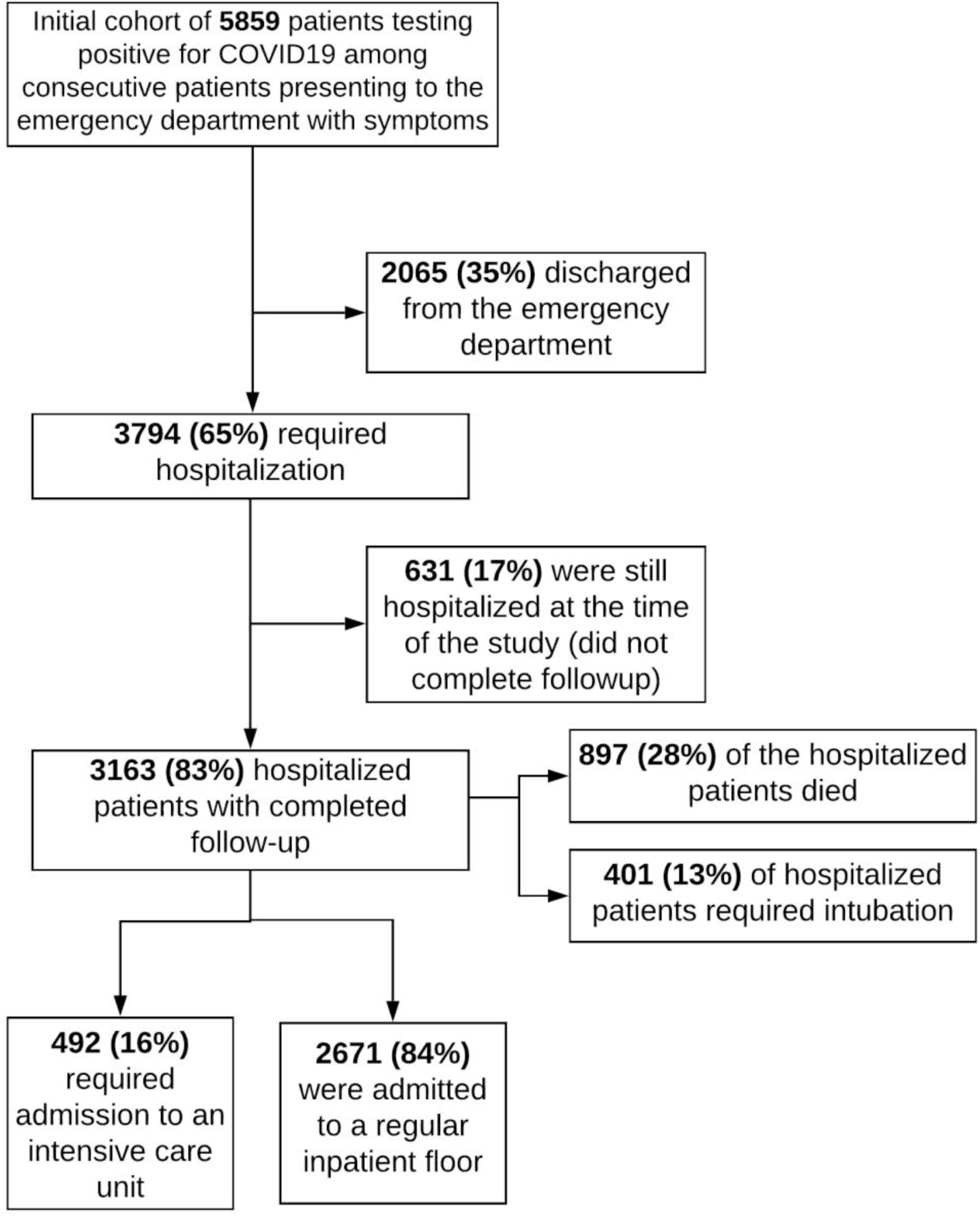
Flow diagram summarizing the course of COVID19-positive patients.

After excluding patients who died within 3 days, the mean LOS was 7.3 days (SD=5.3 days; median=6 days; IQR=6 days), and 2177 of the remaining 2892 patients (75%) required an LOS exceeding 3 days. The characteristics of hospitalized patients are summarized in Figure 3 with comparisons between early- and late-discharge patients. Factors that were independently associated with an extended LOS included older age (OR=1.03 [1.02-1.04]), chronic kidney disease (OR=1.91 [1.35-2.71]), elevated maximum temperature (OR=2.91 [2.40-3.53]), and low minimum percent oxygen saturation (OR=3.89 [3.16-4.79]). The univariable analysis and optimal adjusted model are summarized in Table 5. Age provided better discrimination when employed as a continuous variable. The stepwise model provided an AUC of 0.770 (95% CI: 0.752-0.789). Bootstrap validation yielded a negligible optimism of 0.0029 which translates into a bias-corrected AUC of 0.768. The Hosmer-Lemeshow test yielded a non-significant p-value indicating appropriate calibration.

**Table 5.**
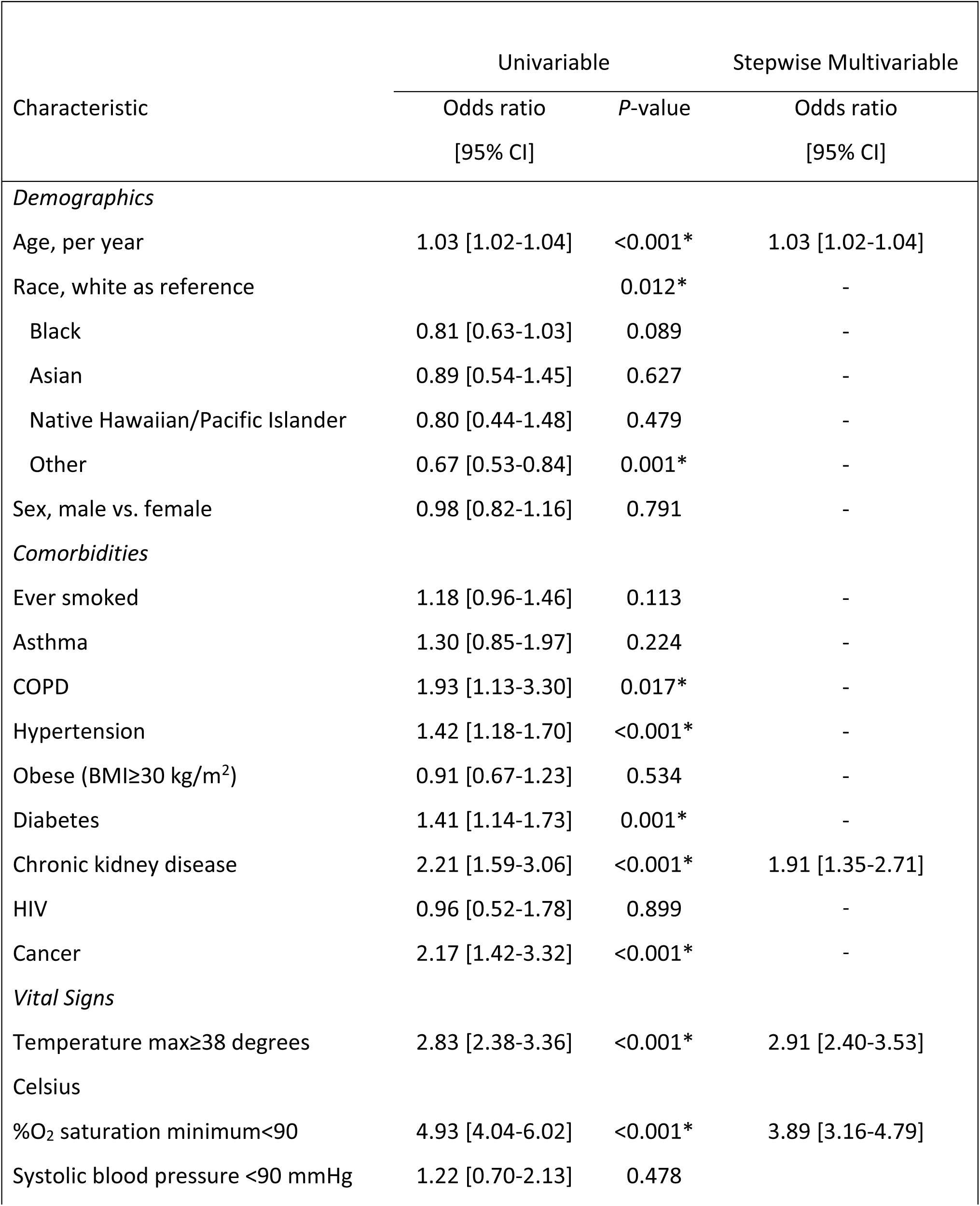
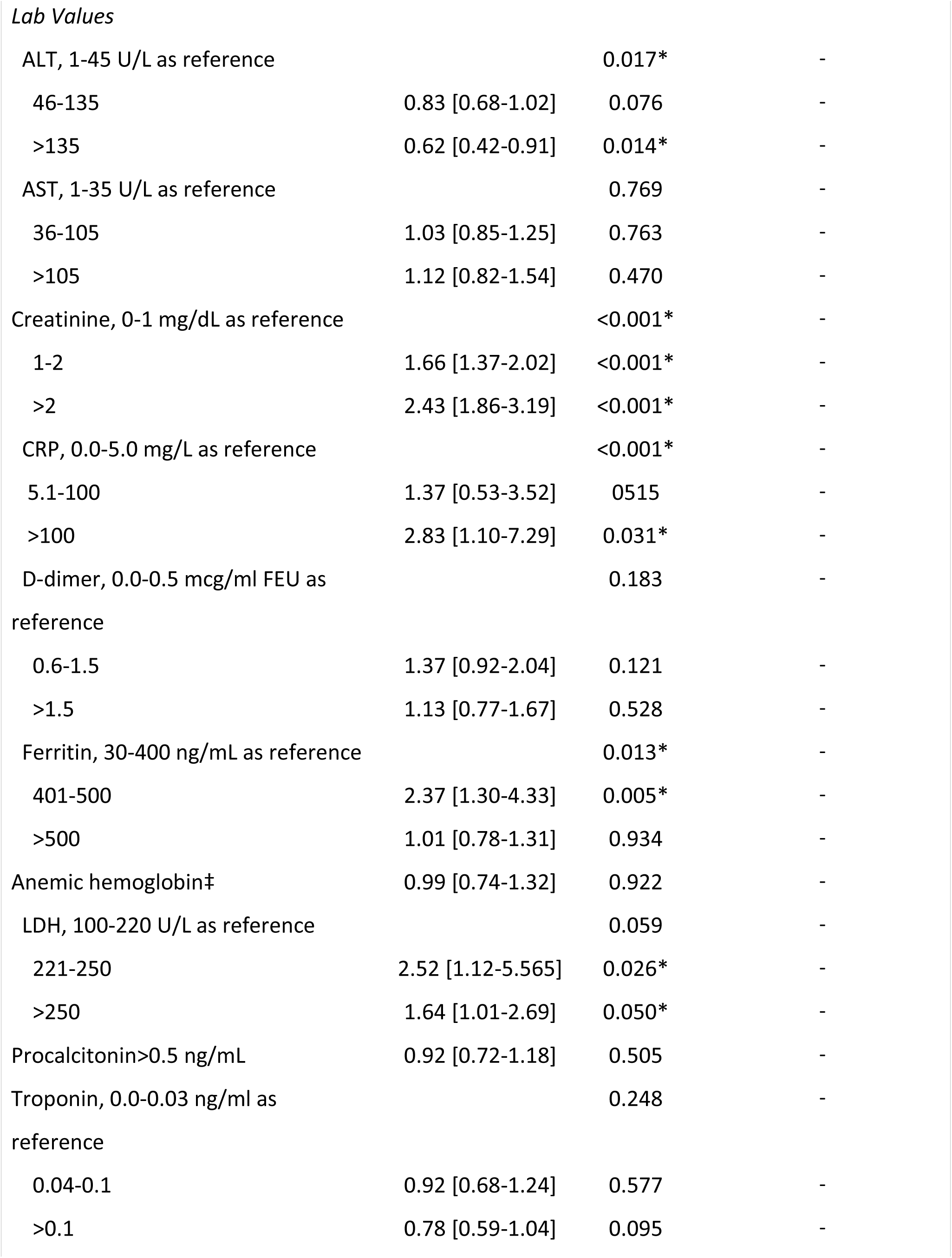
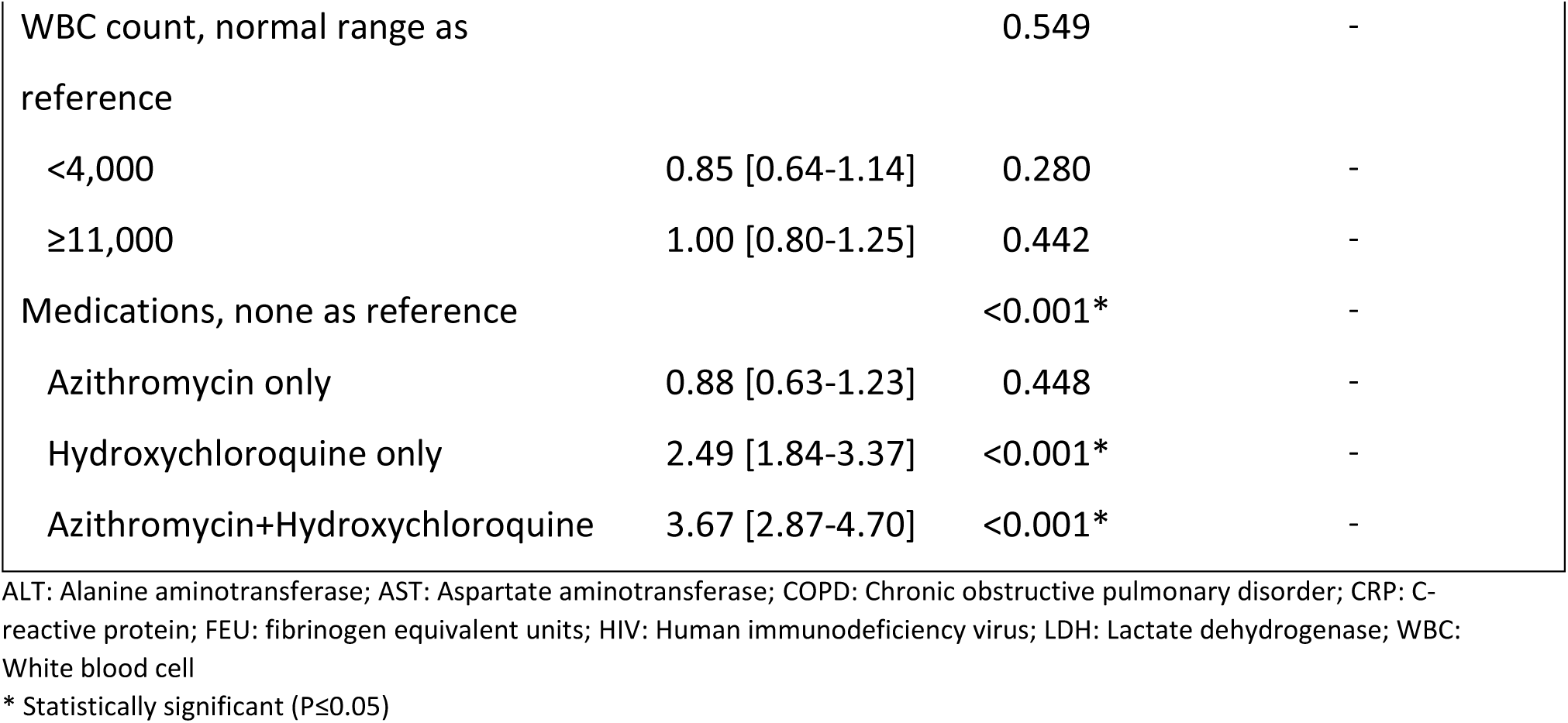
Univariable and multivariable analysis of factors associated with prolonged length of stay (> 3 days) in surviving hospitalized patients (n=2892).

**Supplementary Index: Figure 3.**
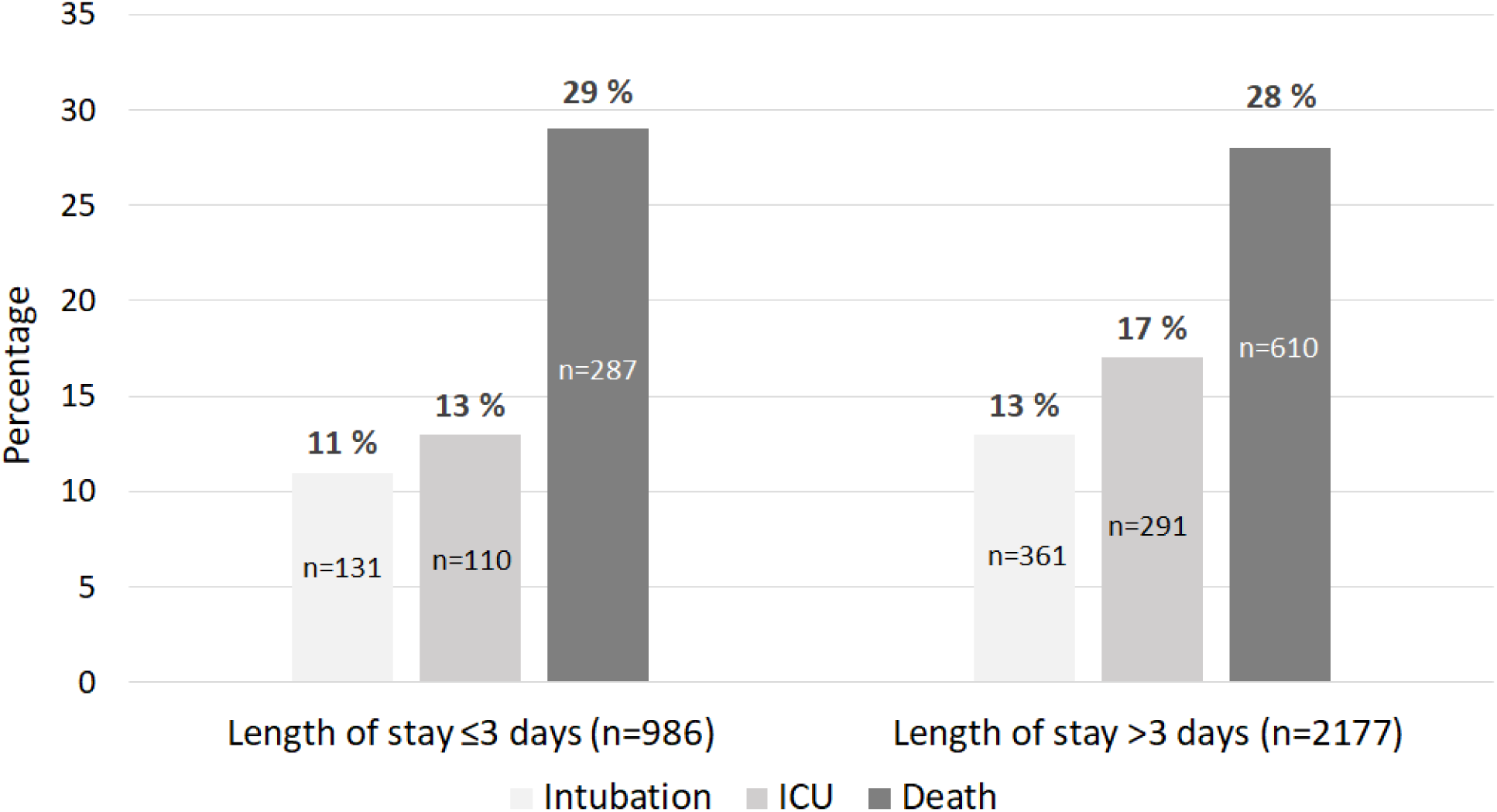
Description of hospitalized patients (n=3163).

A combined calculator of likelihood of hospitalization and extended LOS can be accessed with the following URL:

https://covid19-outcome-prediction.shinyapps.io/COVID19_Hospitalization_Calculator/

## Discussion

We analyzed a large cohort of 5,859 Covid-19 patients in the United States. The results yielded internally validated models with good discrimination that were able to predict both the need for hospitalization in COVID-19 patients presenting to the ED, as well as the risk of prolonged hospitalization among admitted patients. Contingency plans for ongoing secondary COVID-19 resurgences throughout the world will include defining which patients will require acute hospital and prolonged admission.

### Hospitalization

There remains a relative paucity of US studies analyzing hospitalization rates and their determinants, which highlights the gap in knowledge about current management practices. Petrilli et al. analyzed factors associated with hospitalization in a cohort of 4103 COVID-19 patients presenting to New York University (NYU) Langone Health system.^16^ The inclusion criteria however included both inpatient and outpatient visits rather than ED visits; hence, a lower hospitalization rate of 48.7% was reported,^16^ compared to 65% in our cohort which only included patients evaluated in the ED. In concordance with our results, identified risk factors for hospitalization included increased age, chronic kidney disease, diabetes, and male gender. Other important factors included obesity, race, heart failure, hyperlipidemia, and tobacco use.^16^ Similarly, Richardson et al. identified the characteristics of 5700 hospitalized COVID-19 patients at the Northwell Health System, but also excluded ED visits.^17^ Although the authors did not construct predictive risk models in these studies, we expect our calculator to be generalizable to these cohorts and other urban settings given the significant overlap in patient demographics and risk factors, but external validation remains warranted. Given that a large part of the NYU cohort presented to outpatient ambulatory care, vital signs and laboratory studies could not be obtained and analyzed in a large number of patients. Our data revealed that fever and oxygen desaturation are key vital signs associated with illness requiring hospitalization. Important laboratory values that predicted need for hospitalization included AST, ferritin, CRP, and creatinine but not procalcitonin and white blood cell counts. While admission risk calculators have been developed to estimate hospital utilization for COVID-19, they have limited applications for the US adult population since they included pediatric^18^ and/or non-US^19^ populations. Jehi et al. relied on a development cohort of 2852 patients with positive COVID-19 testing in the Cleveland Clinic health system to construct a predictive model for hospitalization which included demographic, comorbidity, and clinical symptom predictors.^20^ The model achieved good discrimination but relied on more than 20 variables and lacked important objective predictors (e.g., vital signs) that are typically available by the time a patient receives a positive test for COVID-19. The advantages of our study lie in the utilization of an exceptionally large multi-ethnic New York population at the height of the first COVID-19 pandemic to develop a concise and practical tool with objective measures of hospitalization risk.

### Length of Stay

One of the largest published studies on hospitalized COVID-19 patients in North America consists of a descriptive analysis of 5700 patients presenting to the Northwell hospital system in New York City.^10^ The reported overall median LOS was 4.1 days, including patients who died during hospitalization. It should be noted however, that 3066 out of the 5700 (54%) patients were still hospitalized at the time of analysis and therefore did not complete follow-up. Among hospitalized patients discharged alive, the median LOS increased from 2.5 days in the 20-29 years age group to 4.8 day in the 90 years and above group.^10^ Although no statistical significance was investigated, this trend closely matches our identified association of extended LOS with increasing age. Rees et al. conducted a systematic review of studies reporting summary measures for hospital length of stay worldwide. The authors reported a pooled median length of stay of 14 days in China and of 5 days outside of China and attributed this difference to variation in criteria for admission and discharge as well as heterogeneity in timing in relation to the pandemic.^21^ Our median length of stay of 6 days is in close agreement with the pooled median of 5 days reported in studies outside of China. Worldwide variation also underscores the importance of verifying applicability of prediction models before implementation across geographical regions. To our knowledge, no internally validated predictive models for LOS derived from readily available demographic, comorbidity, and clinical data in COVID-19 patients exists in the literature. Our data highlights the key predictive value of age, chronic kidney disease, temperature, and oxygen saturation, which can identify patients requiring more hospital resources. Elevated levels of CRP, creatinine, and ferritin also appear to be key determinants of LOS and worth investigating in future studies.

### Mortality

Among 3163 hospitalized patients who completed follow-up, the mortality rate was 28% which is comparable to the rate of 21% identified in the corresponding New York City cohort of 2634 patients.^10^ (Northwell, not NYU) A study by Zhou et al. of 191 hospitalized patients in Wuhan, China similarly reported a mortality rate of 28% (54/191). Our data confirms previously reported mortality rates among hospitalized COVID-19 patients and enhances precision by using the largest hospitalized cohort with completed follow-up to date. Grasselli et al. reported a mortality rate of 26% among 1581 ICU patients with COVID-19 in the Lombardy region of Italy.^6^ In general, case fatality rates reported in the literature include 2.3% (1,023 deaths of 44,672 confirmed cases) in China^22^ and 7.2% (100 out of 1,625 patients) in Italy.^23^ Our ICU admission rate of 16% and intubation rate of 13% mirrored those of Richardson et al. reporting rates of 14% and 12%, respectively in the Northwell cohort.^10^

### Previous Models

A systematic review by Wynants et al., which discussed 10 previously published prognostic models for COVID-19 patients, identified 9 studies: 6 focused on mortality, 2 focused on the development of critical illness, and 1 focused on LOS.^3^ All studies were based in China, and the largest sample size for model derivation was 577 patients. The LOS model was based solely on CT imaging findings and predicted length of stay greater than 10 days using a limited sample of 26 patients with COVID-19 penumonia.^24^ The findings described in our study underscore the importance of incorporating comorbidity and clinical data, which even alone, can explain a large proportion of variance in outcomes. Otherwise, age, sex, hypertension, LDH, and CRP constituted some of the main predictors of mortality and critical illness models.^25–28^ Main criticisms of previous models included the lack of calibration assessment and the absence of a readily available format for use in clinical practice.^3^ Our analysis ensured adequate calibration and rendered the models easily accessible through a user-friendly web-based calculator.

### Utility

The main strength of this calculator lies in its ability to provide accurate discrimination of illness requiring hospitalization and prolonged length of stay based on simple variables that are readily available at the earliest point of contact when a patient presents to an ED. Age, comorbidities, and vital signs can be obtained in a timely fashion by triage personnel. In times of resource shortages, overwhelming disease incidence, and high admission rates, such supplementary tools can help guide the prioritization of patients for subsequent investigations, including laboratory studies and imaging. It can also offer ED physicians a validated tool that facilities management practices in large urban referral centers. The calculator may prove valuable in the absence of clear disseminated guidelines, where there may be uncertainity with regards to increasing the intensity of management. Patients and families additionally inquire about LOS, particularly given increased anxiety from additional exposure to COVID-19 in the hospital setting. This tool might help provide personalized expectations for patients and their families. On a larger scale, the ability to predict length of stay can help hospital systems optimize their allocation of resources and can help gauge capacity to handle additional incoming patients. External validation in the outpatient setting might also provide clinicians with a decision-support tool to refer patients for emergency care. These benefits become highly relevant should further waves of disease occur after the suspension of lockdown.

### Limitations

The findings of this study must be interpreted with caution. This analysis is based on a retrospective analysis of patients presenting to a single hospital health system in New York City, which has been at the epicenter for COVID-19. Our findings might be best extrapolated for use in urban areas with a similar elevated disease burden where the online calculator may be most useful. The concordance of our descriptive data with other published data on COVID-19 increases the confidence in our results but does not eliminate selection bias entirely. Hence, external validation remains warranted. The data lacked specific symptom variables including cough, dyspnea, and pharyngitis but incorporates objective data including vital signs and oxygen desaturation. The calculator is not meant to offer a definitive answer to the management of every COVID-19 patient but can be used to serve as an adjunct to clinical judgement in an ED setting. Given a large amount of missing data in lab values, these were not used to develop the predictive models. Still, clinical data achieved significant discrimination.

## Conclusion

Age, comorbidities and vital signs on admission constitute robust predictors for the need of hospitalization and length of stay in COVID-19 patients presenting to the ED. The prediction tool derived from this study can help design resource allocation during a surge of COVID-19 patients presenting to hospital EDs, help guide quality of care, and assist in designing future studies on the triage and management of patients with COVID-19.

## Data Availability

All data is available upon request

## Declarations

### Conflicts of interest relevant to this work

None

## Acknowledgement

NA.

